# MRI-Targeted Prostate Biopsy Introduces Grade Inflation and Overtreatment

**DOI:** 10.1101/2024.01.10.24300922

**Authors:** Abderrahim Oussama Batouche, Eugen Czeizler, Timo-Pekka Lehto, Andrew Erickson, Tolou Shadbahr, Teemu Daniel Laajala, Joona Pohjonen, Andrew Julian Vickers, Tuomas Mirtti, Antti Sakari Rannikko

## Abstract

**Purpose:** The use of MRI-targeted biopsies has led to lower detection of Gleason Grade Group 1 (GG1) prostate cancer and increased detection of GG2 disease. Although this finding is generally attributed to improved sensitivity and specificity of MRI for aggressive cancers, it might also be explained by grade inflation. Our objective was to determine the likelihood of definitive treatment and risk of post-treatment recurrence for patients with GG2 cancer diagnosed using targeted biopsies relative to men with GG1 cancer diagnosed using systematic biopsies.

**Methods:** We performed a retrospective study on a large tertiary centre registry (HUS Acamedic Datalake) to retrieve data on prostate cancer diagnosis, treatment, and cancer recurrence. We included patients with either GG1 with systematic biopsies (3317 men) or GG2 with targeted biopsies (554 men) from 1993 to 2019. We assessed the risk of curative treatment and recurrence after treatment. Kaplan-Meier survival curves were computed to assess treatment- and recurrence-free survival. Cox proportional hazards regression analysis was performed to assess the risk of posttreatment recurrence.

**Results:** Patients with systematic biopsy detected GG1 cancer had a significantly longer median time-to-treatment (31 months) than those with targeted biopsy detected GG2 cancer (4 months, p<0.0001). The risk of recurrence after curative treatment was similar between groups with the upper bound of 95% CI, excluding an important difference (HR: 0.94, 95% CI [0.71-1.25], p=0.7).

**Conclusion:** GG2 cancers detected by MRI-targeted biopsy are treated more aggressively than GG1 cancers detected by systematic biopsy, despite having similar oncologic risk. To prevent further overtreatment related to the MRI pathway, treatment guidelines from the pre-MRI era need to be updated to consider changes in the diagnostic pathway.

## 1 Introduction

Before the early 2010s, PCa was typically diagnosed using systematic transrectal ultrasound (TRUS)-guided biopsy[1]. Recently, MRI-guided targeted biopsies have gained popularity[2]. MRI targeting has been purported to improve both the sensitivity and specificity of biopsy, avoiding biopsy in men at low risk of high-grade cancer, thereby reducing the risk of overdiagnosis and biopsy-related complications [3, 4], and ensuring that regions of the prostate most likely to harbour aggressive disease are sampled. Currently, clinical guidelines, including those of the European Association of Urology, recommend routine use of MRI before diagnostic prostate biopsy.

There is compelling evidence that targeted biopsy has decreased the detection of low-grade cancers and increased the detection of higher-grade cancers compared to systematic biopsy[3, 5–7]. For instance, in the PRECISION trial, men randomised to the MRI pathway had a 13% absolute decrease in Gleason grade group 1 (GG1) cancers and a 12% increase in GG2 cancers[5]. However, it is plausible that this finding is due, at least in part, to the reclassification of GG1 cancers into GG2, that is, grade inflation. Indirect evidence for grade inflation comes from the observation that MRI-targeting finds many high-grade cancers in groups of men who are known to have a very low risk of prostate cancer mortality, such as those with negative systematic biopsy[8]. However, there have been few direct comparisons of the relative oncologic risk of tumours identified using MRI-targeted versus systematic biopsy.

In this study, we examine large-scale registry data to compare prostate cancer risks by method of cancer detection. Specifically, we compared the recurrence rates between systematic biopsy-detected GG1 prostate cancer and MRI-targeted biopsy-detected GG2 prostate cancer as a direct evaluation of the grade inflation hypothesis.

## 2 Materials and methods

We used the Finnprostate dataset, which is a large patient registry study combining Finnish national healthcare data with local hospital data (n=700 000) of men suspected of having PCa (PSA measured) or diagnosed with PCa (Supplementary Fig. 1). From Finnprostate, we gathered a HUS (Hospital District of Helsinki and Uusimaa) sub-cohort of men (n=326 796) with comprehensive patient information regarding outpatient clinic and hospital visits as well as data regarding laboratory tests, medication, radiological, pathological, and surgical reports, as well as comorbidities from 1993 to 2019. The above data is embedded within the regional HUS Acamedic datalake.

We identified patients with an initial diagnosis of GG1 prostate cancer in systematic biopsies or GG2 prostate cancer in MRI-targeted biopsies. To mitigate the effect of any missing treatment information in our registry on treatments with curative intent, a PSA drop of at least 75% but not less than 3 ng/ml within one year, or at least 50% but not less than 4 ng/ml within one year (this represents 6% of curative treatment data), were considered as an indication of treatment with curative intent, as opposed to active surveillance defined as no curative treatment received or the period from diagnosis until the first curative treatment received. These criteria have been verified to correctly detect 90% of our known first curative treatment data[9]. Depending on the kinetics of the PSA drop, we classified a drop below 0.1ng/ml as radical prostatectomy (RP); otherwise, it was classified as radiation therapy (RT). Sankey diagrams were generated to display the treatment trajectories.

Patients were considered to have experienced recurrence (CR) based on either biochemical recurrence or second-line treatment (Supplementary Fig. 2). Thus, CR considers the clinical reality that some men are being referred to second-line treatment before the official definition of biochemical recurrence has been reached. Biochemical recurrence was defined as a PSA increase over 0.2 ng/ml after RP or 2 ng/ml over the nadir after RT.

### 2.1 Statistical Methods

Our null hypothesis was that there would be no significant difference in risk for recurrence after treatment between GG1 with systematic biopsy and GG2 with MRI-targeted biopsy. Descriptive statistics were used to summarize and present baseline characteristics, while inferential statistics were used to assess the likelihood of definitive treatment and risk of post-treatment recurrence. Kaplan-Meier survival curves and log-rank tests were used to analyze treatment- and relapse-free survival. All analyses were performed using R Statistical Software (v4.1.2; R Core Team 2021), including ‘survival’ and ‘survminer’ packages in R software (version 4.1), while Python (version 3.9) was used to pre-process, clean, and combine the data.

We performed a series of sensitivity analyses to assess the robustness of the conclusions. Relapse-free survival was analyzed in patients treated with RP alone to mitigate the effect of androgen deprivation therapy (ADT) with RT may have had. Men who were eventually treated after a period of surveillance longer than one year were more likely to have their PCa evolved over their initial diagnosed GG. Therefore, we separately analyzed a cohort of patients treated within one year of diagnosis, those treated with RP only within one year of diagnosis, and those diagnosed with GG1 and never upgraded in subsequent biopsies. Based on these four sub-cohorts, we performed Cox proportional hazards regression analysis to assess the risk of post-treatment recurrence in GG1 NoMRI and GG2 MRI patients. Furthermore, these analyses were controlled for PSA levels and the number of positive cores.

## 3 Results

We identified 8407 patients diagnosed with GG1 or GG2 prostate cancer. We excluded patients for whom we were unable to identify their first biopsy with a prostate cancer diagnosis, as well as patients who were over 80 years of age at the time of their first biopsy (Supplementary Fig. 1). The final study cohort consists of 3317 patients with systematic biopsy diagnosed with GG1 cancer and 554 patients with targeted biopsy diagnosed with GG2 cancer (Table 1).

**Table 1.** Demographics of the study cohorts. Clinical recurrence (CR)= Biochemical recurrence event or a secondary treatment; GG= Gleason grade group; MRI= magnetic resonance imaging.

|  | GG1 no MRI | GG2 MRI |
| --- | --- | --- |
| Number of patients | 3317 | 554 |
| Median age at diagnosis, yr (quartiles) | 66 (61-72) | 67 (62-72) |
| Median PSA at diagnosis, ng/ml (quartiles) | 7.3 (5.1-10.5) | 8.1 (5.7-12.7) |
| Clinical recurrence events (CR in 5yr) | 615 (427) | 569 (394) |
| Treatments |  |  |
| Radical prostatectomy | 586 | 165 |
| Radiation therapy | 776 | 104 |
| Medical treatment | 283 | 120 |
|  | (n=1645) | (n=389) |

Treatment patterns were visualised by plotting treatment trajectories in Sankey diagrams for the two groups (Supplementary Fig. 3). In Kaplan-Meier analysis, the likelihood of treatment was significantly lower for men with systematic biopsy detected GG1 compared to men with targeted biopsy detected GG2 HR: 2.77; 95% CI [2.49 – 3.09], p*<*0.0001) (Figure 1). Risk of treatment within one year of diagnosis was 35% for GG1 and 71% for GG2.

**Fig. 1.**
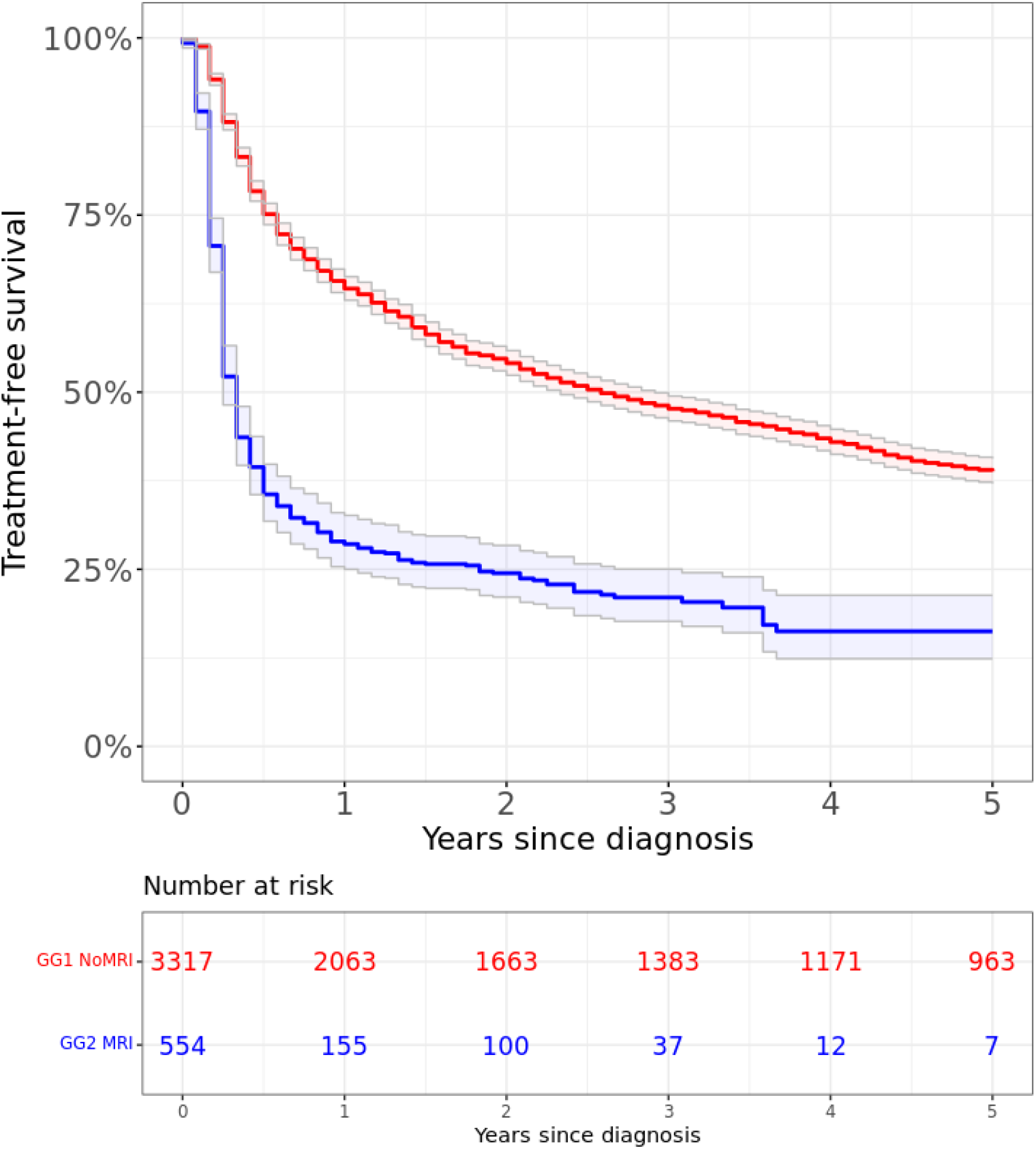
Five-year treatment-free survival (Kaplan-Meier) for GG1 NoMRI (red line) and GG2 MRI (blue line) groups. 95% confidence interval illustrated with corresponding grey lines. The start time is set as the date of diagnosis. HR: 2.77; 95% CI [2.49 – 3.09], Log-rank p*<*0.0001

Next, we compared the pathology at biopsy with that of prostatectomy in RP-treated men. In 75% of the 268 men who underwent targeted biopsy detected GG2 cancer the biopsy pathology was concordant with RP pathology. In 564 men with systematic biopsy-detected GG1 cancer, the concordance was only 33% (p*<*0.0001). In addition, 67% and 16% of the 564 men with GG1 cancer on systematic biopsy and 99% and 24% of the 268 men with GG2 cancer on targeted biopsy, that had radical prostatectomy, had GG2-5 and GG3-5 prostate cancer at final pathology, respectively. Finally, we compared relapse-free survival between men diagnosed with GG1 cancer using systematic biopsy and men diagnosed with GG2 cancer using targeted biopsy. In the Kaplan-Meier plot, the curves virtually overlapped, with no statistically significant difference in median survival (HR: 0.94, 95% CI [0.71-1.25], p=0.7) (Figure 2).

**Fig. 2.**
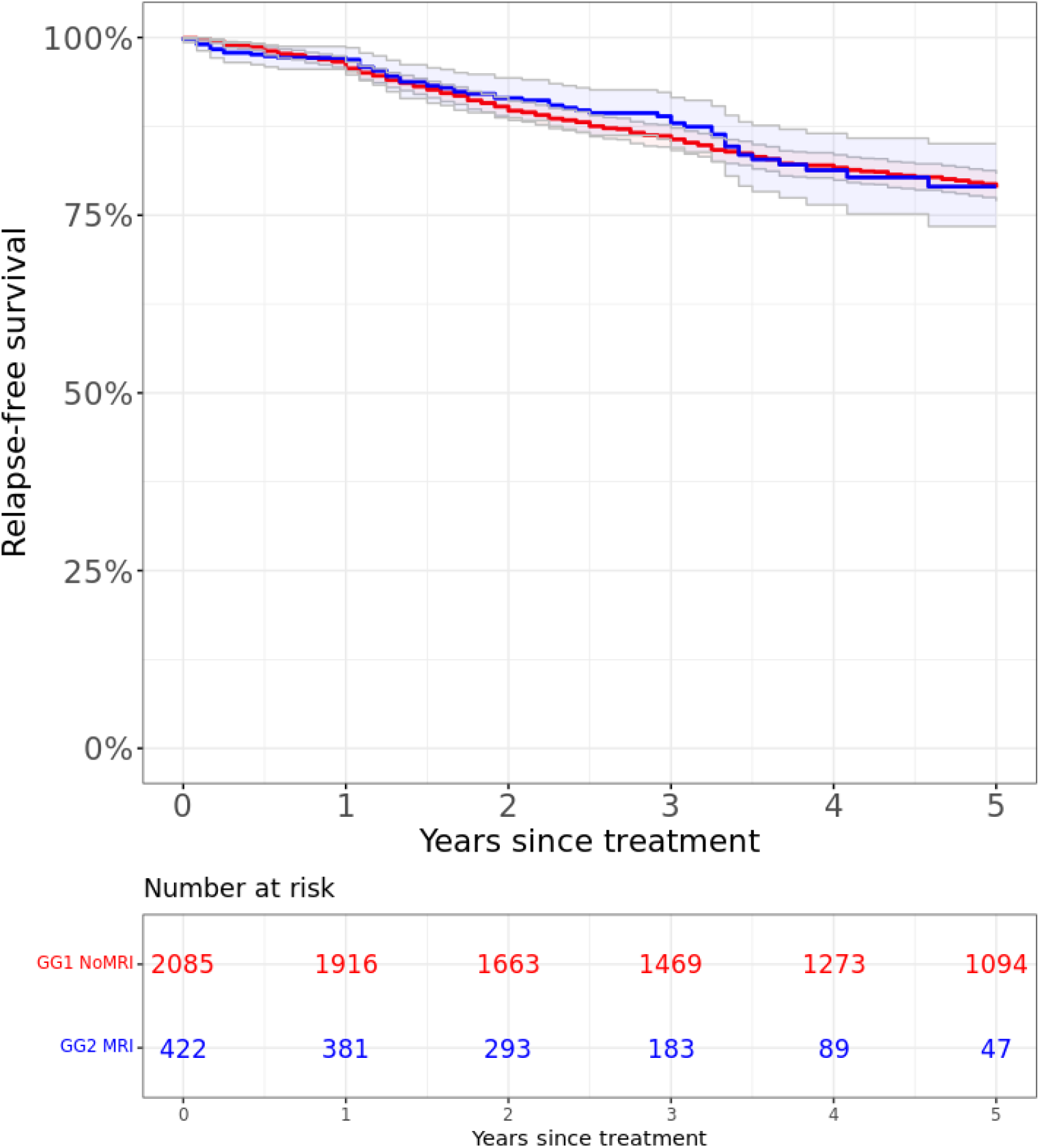
Five-year relapse-free survival (Kaplan-Meier) for GG1 NoMRI (red line) and GG2 MRI (blue line) groups. 95% confidence interval illustrated with corresponding grey lines. The start time is set as the date of treatment. HR=0.94, 95% CI= [0.71-1.25], Log-rank p=0.7.

The results of the sensitivity analyses confirmed the findings of the main analyses. In no analysis was there a statistically significant difference in BCR between systematic GG1 and targeted GG2, and the upper bound of the 95% CI was never greater than 2 (Supplementary Figures 4 – 7 and Supplementary Table 2-3).

## 4 Discussion

The use of MRI-targeted biopsies has led to a lower detection of GG1 prostate cancer and an increased detection of GG2. Although this finding is generally attributed to the improved sensitivity and specificity of MRI for aggressive cancers, it might also be explained by grade inflation. We found direct evidence to support this hypothesis: GG2 cancers detected by MRI-targeted biopsy were not oncologically more aggressive than GG1 cancers detected by systematic biopsy. Not only were differences between groups non-significant, but the upper bound of the 95% CI also excluded a clinically relevant effect. In our main analysis, the upper bound of the hazard ratio between GG2 and GG1 was 1.25, and the highest upper bound found in the multiple sensitivity analyses was 2. Both values were far lower than those typically reported. For instance, in the MSKCC nomogram, the hazard ratio for BCR comparing GG2 to GG1 was approximately 3[10]. Despite similar oncological risks, MRI-detected GG2 cancers were treated more aggressively, clearly suggesting overtreatment.

In current practice, the major distinction is between GG1, which can be managed conservatively, and GG2 or higher, which generally requires treatment. However, a sub-cohort of low-volume pre-MRI era GG ≤ 2 intermediate-risk PCa has been shown to have a similar prognosis to low-risk PCa in AS, according to a recent meta-analysis[11]. Additionally, efforts have been made for more granular risk stratification of intermediate-risk PCa[12, 13]. Furthermore, there is accumulating data on the extent of Gleason pattern 4, total cancer length, PSA density, and perineural invasion determining adverse findings in RP and worse prognosis during follow-up. However, these studies were conducted mainly on systematic biopsy cohorts, and systematic biopsies and targeted biopsies are different and may predict oncologic risk differently[14]. Our results add to the notion that a more precise risk stratification among targeted biopsy-detected cancers is warranted. Hence, before more long-term data accumulate from targeted biopsy studies, treatment decisions should be based on individual risk assessments, not solely on the highest GG stratification and prediction nomograms based on systematic biopsy data[15].

The natural course of the disease is favourable, especially for low- and intermediate-risk PCa, as demonstrated in the recently updated ProtecT trial with only approximately 3% PCa-specific mortality after a 15-year follow-up, irrespective of the treatment[16]. Thus, studies comparing PCa-specific mortality between patients diagnosed with systematic biopsy and targeted biopsy are not expected at any time. Therefore, we considered it justified to use surrogates such as recurrence after curative treatment as an outcome.

The existing literature on targeted biopsy-induced grade inflation is mostly based on data available at diagnosis (pathology at biopsy and radical prostatectomy) and extrapolation into cohorts with virtually no prostate cancer mortality after benign or low-grade prostate cancer on systematic biopsy, even after an extended follow up[8, 17]. While our results and empirical evidence support grade inflation, they are also in line with both the expert opinion raised in the literature[18, 19] and with the available literature findings[14, 20–22]. Vickers 2021[8] addressed the possibility of overtreatment related to the use of targeted biopsies. Combining data on the number of men with high-grade cancers detected only on targeted biopsy with data on death rates in men with benign systematic biopsy findings and taking into account current guidelines recommending treatment for high-grade disease, Vickers found that a very large number of men with MRI-detected cancers would need to be treated in order to prevent one death.

Our results support the findings of two previous studies comparing the pathological concordance of systematic and targeted biopsies to radical prostatectomy[20, 21]. Downgrading to GG1 after radical prostatectomy was uncommon regardless of the biopsy method while upgrading was significantly more common in men with GG1 on systematic biopsy. Thus, many men with GG2 on targeted biopsy would often have GG1 on systematic biopsy and are candidates for active surveillance. However, GG1 patients treated with radical prostatectomy likely represent a bias towards a more aggressive cohort of men than GG1 patients, on average.

Indirect evidence for grade inflation also comes from registry studies in which men with negative systematic biopsies have been followed for decades with negligible PCa-specific mortality[23–25]. Furthermore, there is abundant literature showing that up to 30% of men with clinical suspicion of PCa but with negative systematic biopsy de facto harbour high-grade prostate cancer[26]. If these are clinically significant, one would expect PCa mortality during extended follow-up[23].

A clinical implication of our results in the context of the existing literature is that more men with MRI-era GG2 prostate cancer should undergo active surveillance (AS). Ideally, AS protocols should be individualised based on known risk factors, and men should preferably be included in prospective AS trials such as PRIAS[27] or SPCG-17[28], which should be strongly emphasised in guidelines. In particular, we agree with the statement, “It is not the MRI that is harming by finding indolent grade group 2 cancers that would have been missed by systematic biopsy; the harm is caused by guidelines telling us to treat most of those cancers.”[29, 30].

Our study is prone to some limitations. In particular, one could expect that the subcohort of men with GG1 curatively treated are the ones considered to harbour features of more aggressive disease. Furthermore, a man followed for GG1 cancer may progress to GG2 during follow-up and therefore would be expected to have a similar risk for progression as those with GG2 at diagnosis. Therefore, we performed a series of sensitivity analyses in this study. First, we restricted the analyses to men treated in the first year only to exclude most men who progressed. Next, we excluded men who had progressed on subsequent biopsies from the analyses. Finally, we adjusted the risk of recurrence for PSA level and the number of positive cores as covariates in the multivariable Cox regression model. These sensitivity analyses confirmed our original findings, with no statistically significant differences between groups, and the upper bound of all 95% CI was far below published estimates of the hazard ratio for GG2 vs. GG1 disease. Due to the registry nature of our study, some of the data were granular, and some of the missing data were inferred (e.g., curative treatment based on PSA changes). This could have caused inaccuracies in the results. However, the effect should be limited based on a quality assessment of the data. Clinical indications for MRI and curative treatments are not known, and indications likely have changed during the study period, which represents a limitation, although it is unclear if this would cause a bias. The Gleason grading also changed during the study period. This has caused risk inflation, especially for GG1, with a bias favouring the GG1 group, further strengthening our results and interpretation. The strengths of our study lie in the large sample size and the comprehensive clinical data available for each patient from diagnosis to treatment and follow-up. Our registry included patients from the beginning of the MRI era, allowing for a comparatively long follow-up period. This extended follow-up period provides valuable insights into the long-term effects of treatments or interventions and helps us better understand outcomes over time.

## 5 Conclusion

GG2 cancers detected by MRI-targeted biopsy are treated more aggressively than GG1 cancers detected by systematic biopsy, despite having similar oncologic risks. To prevent further overtreatment related to the MRI pathway, treatment guidelines from the pre-MRI era need to be updated to consider changes in the diagnostic pathway.

## Data Availability

All data produced in the present study are NOT available

## References

[1] Ouzzane, A., Coloby, P., Mignard, J.-P., Allegre, J.-P., Soulie, M., Rebillard, X., Salomon, L., Villers, A., Committee of Infectious Diseases of the French Association of Urology (CIAFU), Committee of Cancerology of the French Association of Urology (CCAFU): [Recommendations for best practice for prostate biopsy]. Prog Urol 21(1), 18–28 (2011) 10.1016/j.purol.2010.07.001

[2] Kaneko, M., Sugano, D., Lebastchi, A.H., Duddalwar, V., Nabhani, J., Haiman, C., Gill, I.S., Cacciamani, G.E., Abreu, A.L.: Techniques and Outcomes of MRI-TRUS Fusion Prostate Biopsy. Curr Urol Rep 22(4), 27 (2021) 10.1007/s11934-021-01037-x

[3] Matulevičcius, A., Bakavičcius, A., Ulys, A., Trakymas, M., Ušsinskieneė, J., Narušsevičiūtė, I., Sabaliauskaitė, R., Žukauskaitė, K., Jarmalaitė, S., Jankevičius, F.: Multiparametric MRI Fusion-Guided Prostate Biopsy for Detection of Clinically Significant Prostate Cancer Eliminates the Systemic Prostate Biopsy. Applied Sciences 12(19), 10151 (2022) 10.3390/app121910151. Accessed 2023-02-23

[4] Wegelin, O., Exterkate, L., Leest, M., Kelder, J.C., Bosch, J.L.H.R., Barentsz, J.O., Somford, D.M., Melick, H.H.E.: Complications and Adverse Events of Three Magnetic Resonance Imaging–based Target Biopsy Techniques in the Diagnosis of Prostate Cancer Among Men with Prior Negative Biopsies: Results from the FUTURE Trial, a Multicentre Randomised Controlled Trial. European Urology Oncology 2(6), 617–624 (2019) 10.1016/j.euo.2019.08.007. Accessed 2023-03-06

[5] Kasivisvanathan, V., Rannikko, A.S., Borghi, M., Panebianco, V., Mynderse, L.A., Vaarala, M.H., Briganti, A., Budäus, L., Hellawell, G., Hindley, R.G., Roobol, M.J., Eggener, S., Ghei, M., Villers, A., Bladou, F., Villeirs, G.M., Virdi, J., Boxler, S., Robert, G., Singh, P.B., Venderink, W., Hadaschik, B.A., Ruffion, A., Hu, J.C., Margolis, D., Crouzet, S., Klotz, L., Taneja, S.S., Pinto, P., Gill, I., Allen, C., Giganti, F., Freeman, A., Morris, S., Punwani, S., Williams, N.R., Brew-Graves, C., Deeks, J., Takwoingi, Y., Emberton, M., Moore, C.M.: MRI-Targeted or Standard Biopsy for Prostate-Cancer Diagnosis. N Engl J Med 378(19), 1767–1777 (2018) 10.1056/NEJMoa1801993. Accessed 2023-03-02

[6] O’Brien, M.F., Cronin, A.M., Fearn, P.A., Smith, B., Stasi, J., Guillonneau, B., Scardino, P.T., Eastham, J.A., Vickers, A.J., Lilja, H.: Pretreatment prostate-specific antigen (PSA) velocity and doubling time are associated with outcome but neither improves prediction of outcome beyond pretreatment PSA alone in patients treated with radical prostatectomy. J Clin Oncol 27(22), 3591–3597 (2009) 10.1200/JCO.2008.19.9794

[7] Ahmed, H.U., El-Shater Bosaily, A., Brown, L.C., Gabe, R., Kaplan, R., Parmar, M.K., Collaco-Moraes, Y., Ward, K., Hindley, R.G., Freeman, A., Kirkham, A.P., Oldroyd, R., Parker, C., Emberton, M.: Diagnostic accuracy of multi-parametric MRI and TRUS biopsy in prostate cancer (PROMIS): a paired validating confirmatory study. The Lancet 389(10071), 815–822 (2017) 10.1016/S0140-6736(16)32401-1. Accessed 2023-03-04

[8] Vickers, A.J.: Effects of Magnetic Resonance Imaging Targeting on Overdiagnosis and Overtreatment of Prostate Cancer. European Urology 80(5), 567–572 (2021) 10.1016/j.eururo.2021.06.026. Accessed 2023-03-06

[9] Batouche, A.O., Czeizler, E., Koskinen, M., Mirtti, T., Rannikko, A.S.: Synergizing Data Imputation and Electronic Health Records for Advancing Prostate Cancer Research: Challenges, and Practical Applications (2023) 10.48550/ARXIV.2311.02086. Publisher: arXiv Version Number: 1. Accessed 2023-11-17

[10] Prostate Cancer Nomograms: Dynamic Prostate Cancer Nomogram: Coefficients | Memorial Sloan Kettering Cancer Center. https://www.mskcc.org/nomograms/prostate/preop/coefficients Accessed 2023-11-29

[11] Baboudjian, M., Breda, A., Rajwa, P., Gallioli, A., Gondran-Tellier, B., Sanguedolce, F., Verri, P., Diana, P., Territo, A., Bastide, C., Spratt, D.E., Loeb, S., Tosoian, J.J., Leapman, M.S., Palou, J., Ploussard, G.: Active Surveillance for Intermediate-risk Prostate Cancer: A Systematic Review, Meta-analysis, and Metaregression. Eur Urol Oncol 5(6), 617–627 (2022) 10.1016/j.euo.2022.07.004

[12] Zumsteg, Z.S., Zelefsky, M.J., Woo, K.M., Spratt, D.E., Kollmeier, M.A., McBride, S., Pei, X., Sandler, H.M., Zhang, Z.: Unification of favourable intermediate-, unfavourable intermediate-, and very high-risk stratification criteria for prostate cancer. BJU Int 120(5B), 87–95 (2017) 10.1111/bju.13903

[13] Kweldam, C.F., Van Leenders, G.J., Van Der Kwast, T.: Grading of prostate cancer: a work in progress. Histopathology 74(1), 146–160 (2019) 10.1111/his.13767. Accessed 2023-11-17

[14] Gaffney, C.D., Tin, A.L., Fainberg, J., Fine, S., Jibara, G., Touijer, K., Eastham, J., Scardino, P., Laudone, V., Vickers, A.J., Ehdaie, B.: The oncologic risk of magnetic resonance imaging-targeted and systematic cores in patients treated with radical prostatectomy. Cancer (2023) 10.1002/cncr.34981

[15] Leenders, G.J.L.H., Kwast, T.H., Grignon, D.J., Evans, A.J., Kristiansen, G., Kweldam, C.F., Litjens, G., McKenney, J.K., Melamed, J., Mottet, N., Paner, G.P., Samaratunga, H., Schoots, I.G., Simko, J.P., Tsuzuki, T., Varma, M., Warren, A.Y., Wheeler, T.M., Williamson, S.R., Iczkowski, K.A., ISUP Grading Workshop Panel Members: The 2019 International Society of Urological Pathology (ISUP) Consensus Conference on Grading of Prostatic Carcinoma. Am J Surg Pathol 44(8), 87–99 (2020) 10.1097/PAS.0000000000001497

[16] Hamdy, F.C., Donovan, J.L., Lane, J.A., Metcalfe, C., Davis, M., Turner, E.L., Martin, R.M., Young, G.J., Walsh, E.I., Bryant, R.J., Bollina, P., Doble, A., Doherty, A., Gillatt, D., Gnanapragasam, V., Hughes, O., Kockelbergh, R., Kynaston, H., Paul, A., Paez, E., Powell, P., Rosario, D.J., Rowe, E., Mason, M., Catto, J.W.F., Peters, T.J., Oxley, J., Williams, N.J., Staffurth, J., Neal, D.E.: Fifteen-Year Outcomes after Monitoring, Surgery, or Radiotherapy for Prostate Cancer. N Engl J Med 388(17), 1547–1558 (2023) 10.1056/NEJMoa2214122. Accessed 2023-05-17

[17] Carlsson, S., Benfante, N., Alvim, R., Sjoberg, D.D., Vickers, A., Reuter, V.E., Fine, S.W., Vargas, H.A., Wiseman, M., Mamoor, M., Ehdaie, B., Laudone, V., Scardino, P., Eastham, J., Touijer, K.: Long-Term Outcomes of Active Surveillance for Prostate Cancer: The Memorial Sloan Kettering Cancer Center Experience. J Urol 203(6), 1122–1127 (2020) 10.1097/JU.0000000000000713

[18] Next generation imaging (NGI) in prostate cancer: scan or not to scan? That is the question. Technical report, Nordisk Urologisk Förening (Scandinavian Association of Urology), Helsinki, Finland (June 2022)

[19] Active surveillance for intermediate risk prostate cancer: What urologists and patients should know. Technical report, Amsterdam, The Nederlands (July 2022)

[20] Ahdoot, M., Wilbur, A.R., Reese, S.E., Lebastchi, A.H., Mehralivand, S., Gomella, P.T., Bloom, J., Gurram, S., Siddiqui, M., Pinsky, P., Parnes, H., Linehan, W.M., Merino, M., Choyke, P.L., Shih, J.H., Turkbey, B., Wood, B.J., Pinto, P.A.: MRI-Targeted, Systematic, and Combined Biopsy for Prostate Cancer Diagnosis. N Engl J Med 382(10), 917–928 (2020) 10.1056/NEJMoa1910038. Accessed 2023-05-24

[21] Martini, A., Touzani, A., Mazzone, E., Roumiguié, M., Marra, G., Valerio, M., Beauval, J.B., Campi, R., Minervini, A., Berg, R.C.N., Soeterik, T.F.W., Zhuang, J., Guo, H., Gontero, P., Montorsi, F., Briganti, A., Gandaglia, G., Ploussard, G., Young Academic Urologists Working Group on Prostate Cancer of the European Association of Urology, Berg, R.C.N.: Overdiagnosis and stage migration of ISUP 2 disease due to mpMRI-targeted biopsy: facts or fictions. Prostate Cancer Prostatic Dis 25(4), 794–796 (2022) 10.1038/s41391-022-00606-6. Accessed 2023-02-23

[22] Mesko, S., Marks, L., Ragab, O., Patel, S., Margolis, D.A., Demanes, D.J., Kamrava, M.: Targeted Prostate Biopsy Gleason Score Heterogeneity and Implications for Risk Stratification. American Journal of Clinical Oncology 41(5), 497–501 (2018) 10.1097/COC.0000000000000308. Accessed 2023-05-20

[23] Stroomberg, H.V., Andersen, M.C.M., Helgstrand, J.T., Larsen, S.B., Vickers, A.J., Brasso, E.K., Røder, A.: Standardized prostate cancer incidence and mortality rates following initial non-malignant biopsy result. BJU International, 15997 (2023) 10.1111/bju.15997. Accessed 2023-05-20

[24] Hilscher, M., Røder, A., Helgstrand, J.T., Klemann, N., Brasso, K., Vickers, A.J., Stroomberg, H.V.: Risk of prostate cancer and death after benign transurethral resection of the prostate—A 20-year population-based analysis. Cancer 128(20), 3674–3680 (2022) 10.1002/cncr.34407. Accessed 2023-05-20

[25] Stroomberg, H.V., Brasso, K., Røder, A.: Fact, overdiagnosis cannot be evaluated by comparing histological grading of prostate biopsy to prostatectomy specimen. Prostate Cancer Prostatic Dis (2022) 10.1038/s41391-022-00629-z. Accessed 2023-05-20

[26] Fütterer, J.J., Briganti, A., De Visschere, P., Emberton, M., Giannarini, G., Kirkham, A., Taneja, S.S., Thoeny, H., Villeirs, G., Villers, A.: Can Clinically Significant Prostate Cancer Be Detected with Multiparametric Magnetic Resonance Imaging? A Systematic Review of the Literature. European Urology 68(6), 1045–1053 (2015) 10.1016/j.eururo.2015.01.013. Accessed 2023-06-29

[27] Bokhorst, L.P., Valdagni, R., Rannikko, A., Kakehi, Y., Pickles, T., Bangma, C.H., Roobol, M.J.: A Decade of Active Surveillance in the PRIAS Study: An Update and Evaluation of the Criteria Used to Recommend a Switch to Active Treatment. European Urology 70(6), 954–960 (2016) 10.1016/j.eururo.2016.06.007. Accessed 2023-06-20

[28] Ahlberg, M.S., Adami, H.-O., Beckmann, K., Bertilsson, H., Bratt, O., Cahill, D., Egevad, L., Garmo, H., Holmberg, L., Johansson, E., Rannikko, A., Van Hemelrijck, M., Jäderling, F., Wassberg, C., Åberg, U.W.N., Bill-Axelson, A.: PCASTt/SPCG-17-a randomised trial of active surveillance in prostate cancer: rationale and design. BMJ Open 9(8), 027860 (2019) 10.1136/bmjopen-2018-027860

[29] Padhani, A.R., Villeirs, G., Ahmed, H.U., Panebianco, V., Schoots, I.G., Tempany, C.M.C., Weinreb, J., Barentsz, J.O.: Platinum Opinion Counterview: The Evidence Base for the Benefit of Magnetic Resonance Imaging-directed Prostate Cancer Diagnosis is Sound. European Urology 78(3), 307–309 (2020) 10.1016/j.eururo.2020.05.038. Accessed 2023-03-06

[30] Bergh, R.C.N., Rouviére, O., Kwast, T., EAU-EANM-ESTRO-ESUR-SIOG Prostate Cancer Guidelines Panel: Re: Andrew Vickers, Sigrid V. Carlsson, Matthew Cooperberg. Routine Use of Magnetic Resonance Imaging for Early Detection of Prostate Cancer Is Not Justified by the Clinical Trial Evidence. Eur Urol 2020;78:304-6: Prebiopsy MRI: Through the Looking Glass. Eur Urol 78(3), 310–313 (2020) 10.1016/j.eururo.2020.06.005

