## Supplementary for "MRI-Targeted Prostate Biopsy Introduces Grade Inflation and Overtreatment"

**Supplementary Materials**

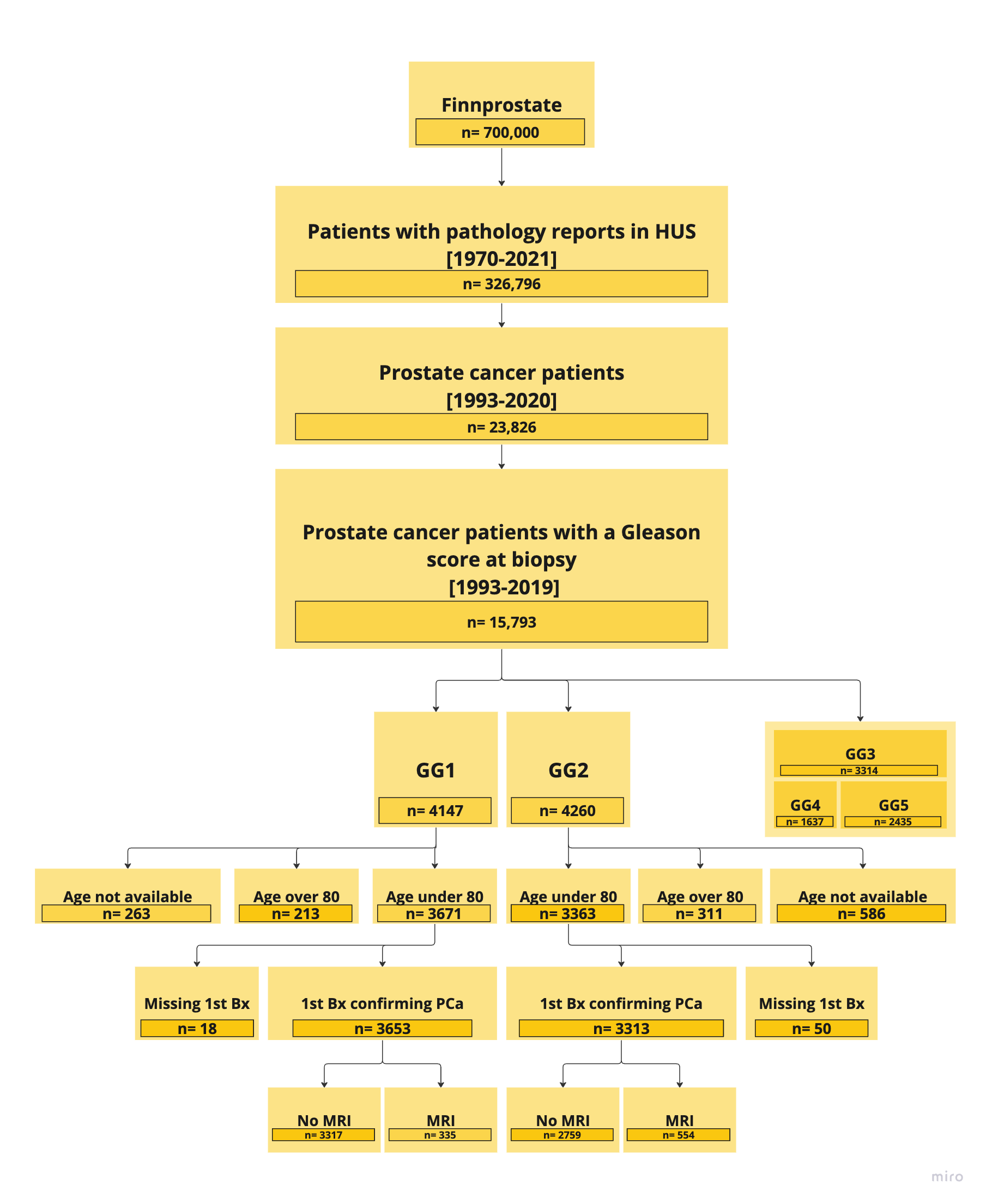

**Supplementary fig. 1**. Flowchart of the patient selection process. HUS=Hospital district of Helsinki and Uusimaa; GG=Gleason grade group; Bx=biopsy; MRI=magnetic resonance imaging.

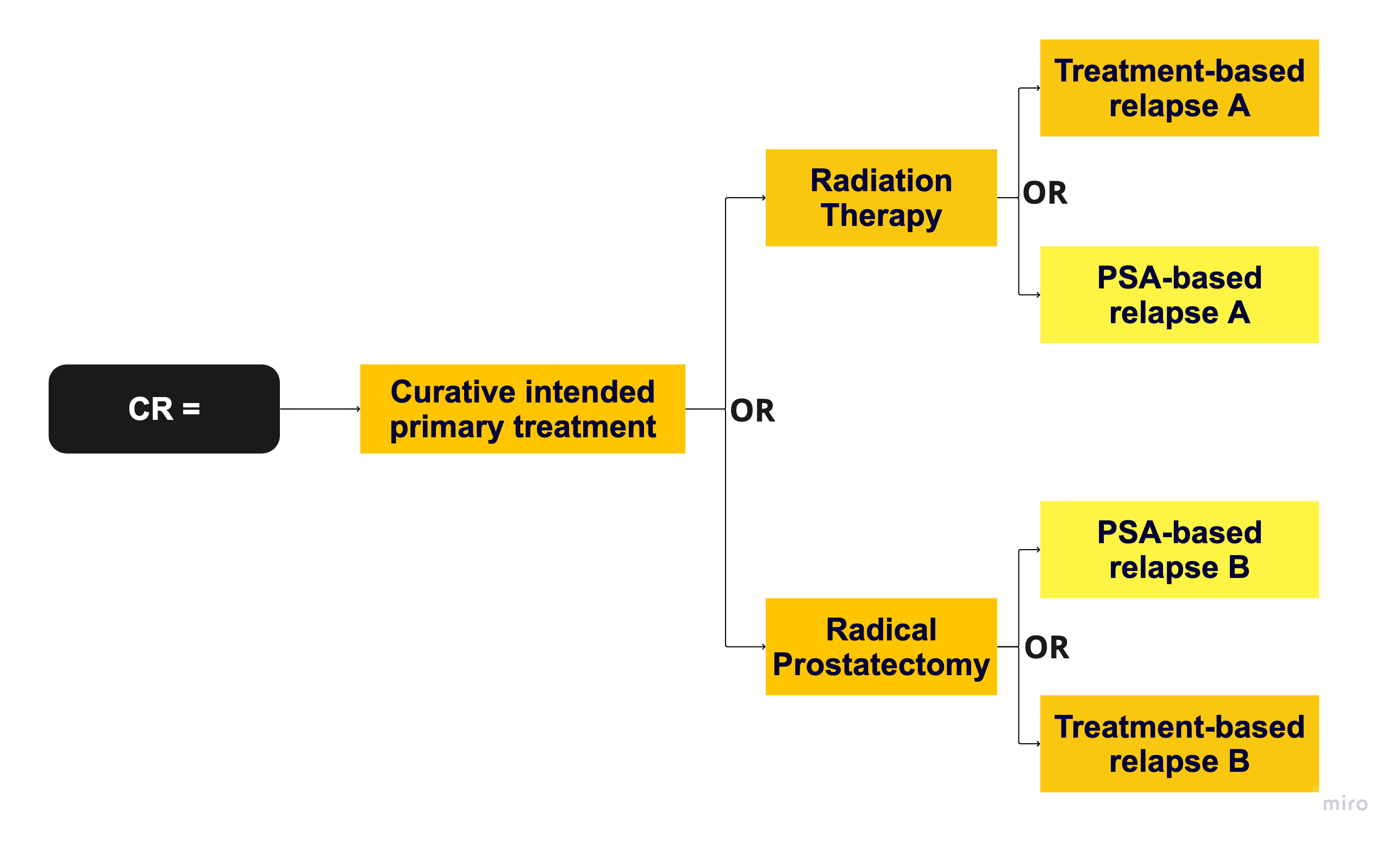

**Supplementary fig. 2**. Clinical relapse (CR) definition. CR is defined as the earliest date of either biochemical recurrence or commencement of secondary therapies after primary treatment. PSA=Prostate specific antigen.

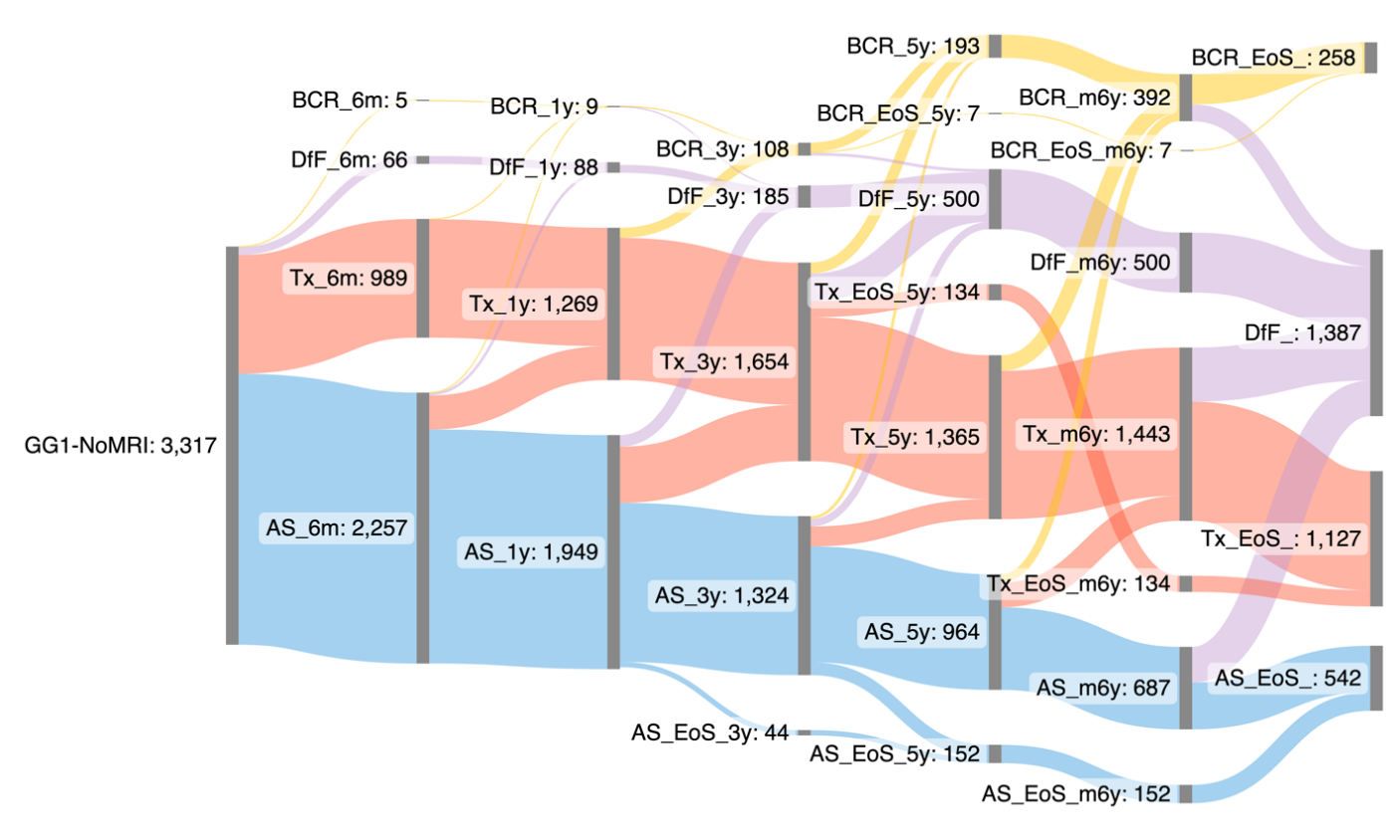

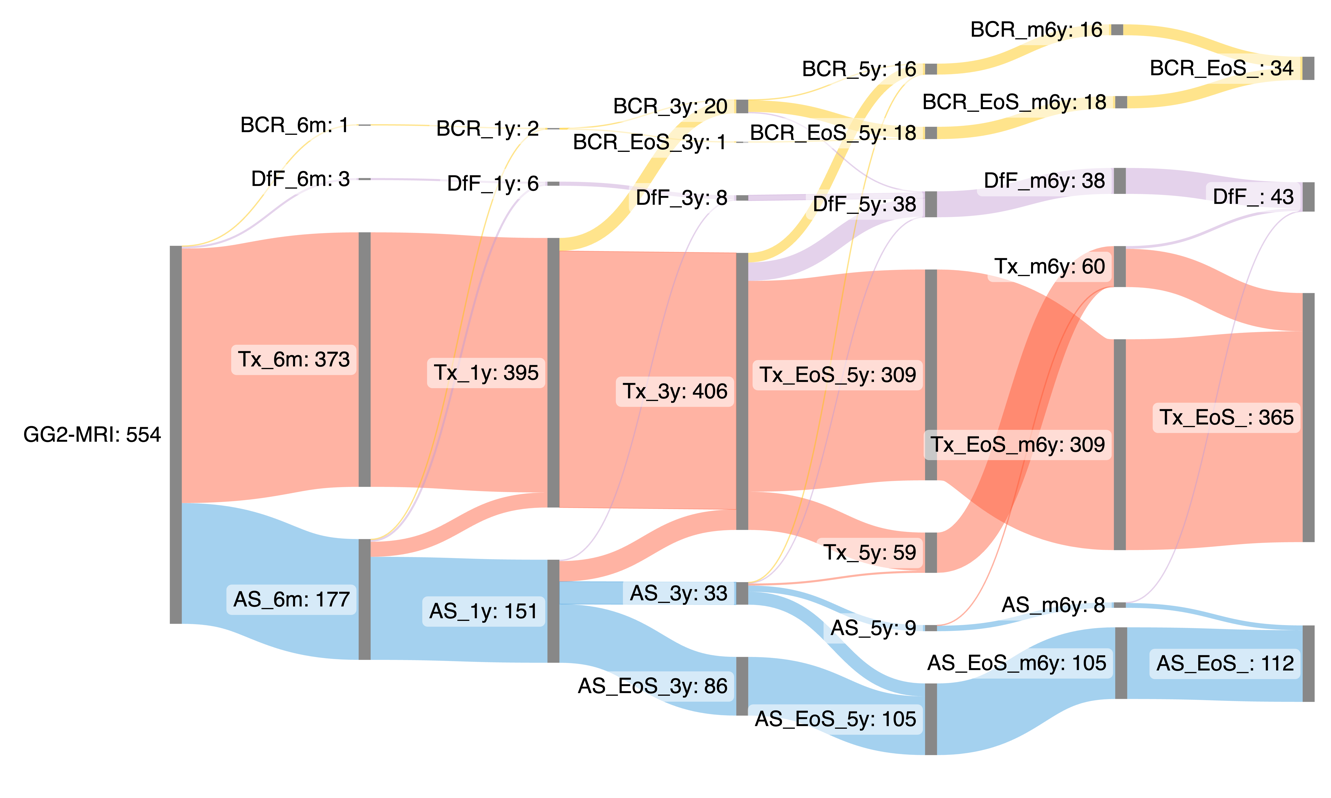

**Supplementary fig. 3.** Sankey diagrams depicting treatment trajectories in the two groups. No curative treatments given in blue, first curative treatment in red.

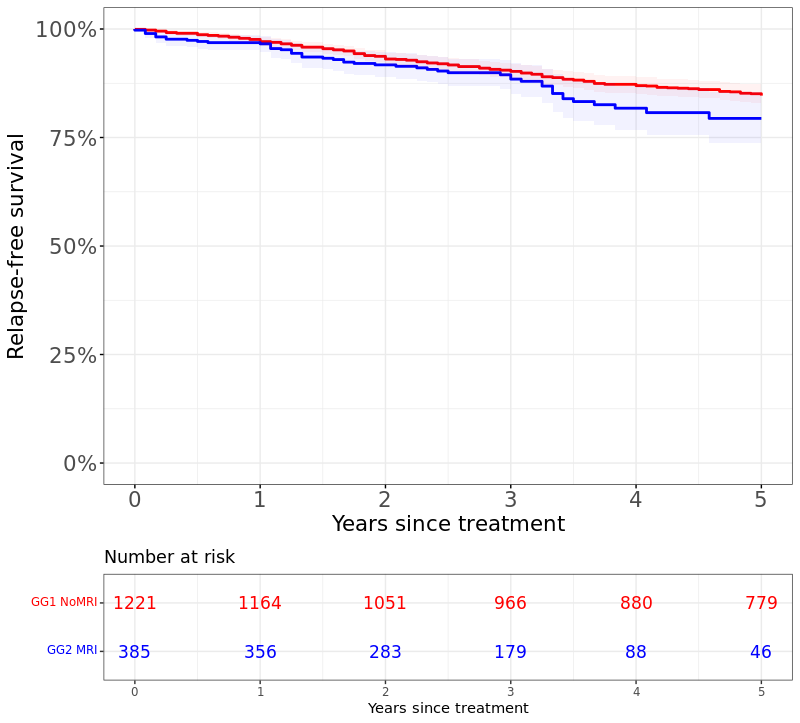

**Supplementary fig. 4.** Five-year relapse-free survival (Kaplan-Meier) for GG1 NoMRI (red line) and GG2 MRI (blue line) groups treated within the first year from diagnosis. 95% confidence interval illustrated with corresponding grey lines. The start time is set as the date of the first curative treatment. HR=1.36, 95% CI= [0.99-1.87], Log-rank p=0.056

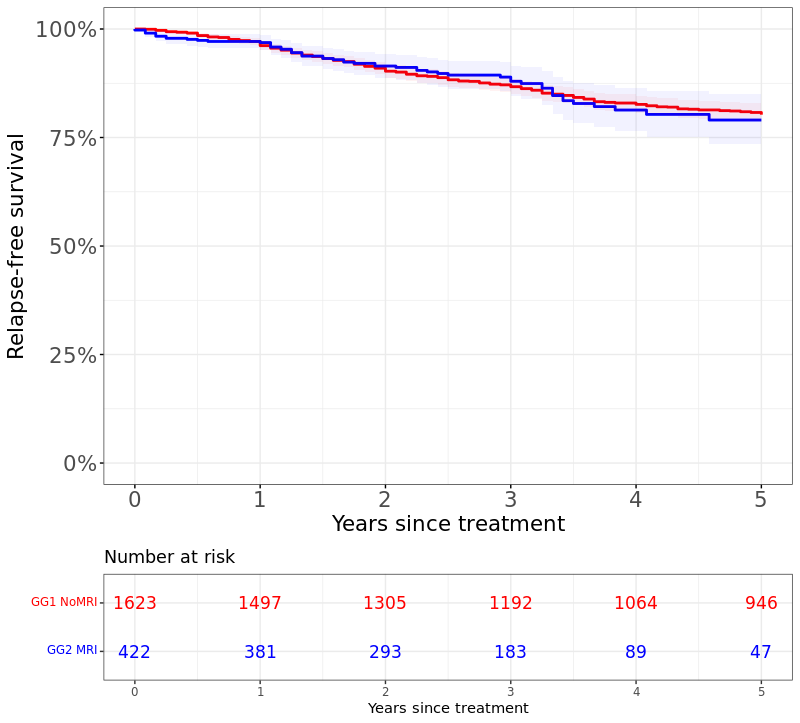

**Supplementary fig. 5.** Five-year relapse-free survival (Kaplan-Meier) for GG1 NoMRI not upgrading on subsequent biopsy (red line) and GG2 MRI (blue line) groups. 95% confidence interval illustrated with corresponding grey lines. The start time is set as the date of the first curative treatment. HR=0.95, 95% CI= [0.76-1.36], Log-rank p=0.9

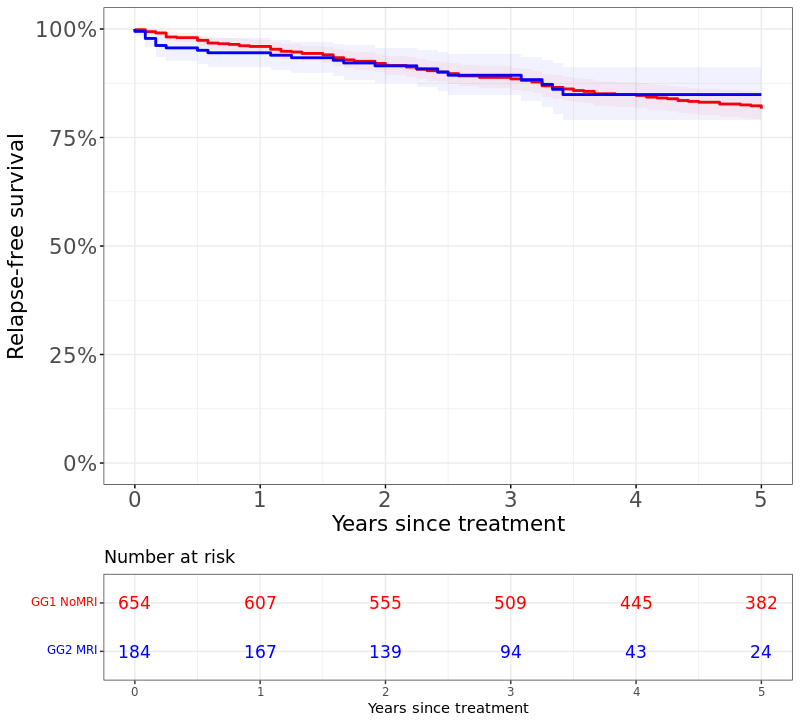

**Supplementary fig. 6.** Five-year relapse-free survival (Kaplan-Meier) for GG1 NoMRI (red line) and GG2 MRI (blue line) groups treated with RP. 95% confidence interval illustrated with corresponding grey lines. The start time is set as the date of the first curative treatment. HR=0.96, 95% CI= [0.61-1.53], Log-rank p=0.9

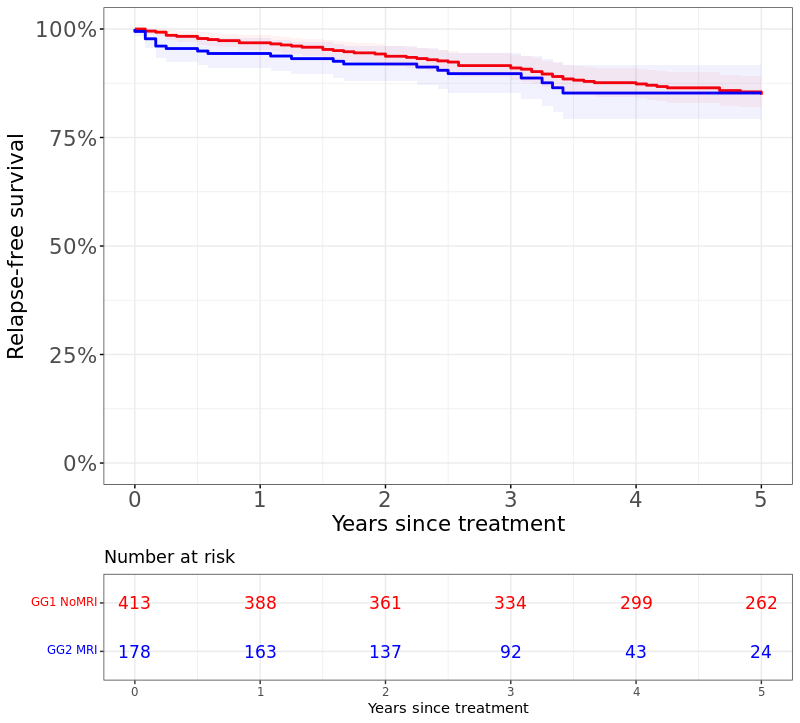

**Supplementary fig. 7.** Five-year relapse-free survival (Kaplan-Meier) for GG1 NoMRI (red line) and GG2 MRI (blue line) groups treated with RP within the first year from diagnosis. 95% confidence interval illustrated with corresponding grey lines. The start time is set as the date of the first curative treatment. HR=1.18, 95% CI= [0.71-1.95], Log-rank p=0.5

| **Cohorts** | **Filter** | **INR (C1/C2)** | **FNR (C1/C2)** | **HR** | **95% CI** | **P-value** |
| --- | --- | --- | --- | --- | --- | --- |
| C1. GG1-NoMRI C2. GG2-MRI | No filter (Fig 2.) | 2085/422 | 1094/47 | 0.94 | 0.71-1.3 | 0.7 |
|  | Treated within 1yr | 1221/385 | 779/46 | 1.36 | 0.99-1.87 | 0.056 |
|  | GG1 Not upgrading on subsequent biopsy | 1623/422 | 946/47 | 0.95 | 0.76-1.36 | 0.9 |
|  | Treated with RP | 654/184 | 382/24 | 0.96 | 0.61-1.53 | 0.9 |
|  | Treated with RP within 1yr | 413/178 | 262/24 | 1.18 | 0.71-1.95 | 0.5 |

**Supplementary table 2.** Results of the sensitivity analysis of the recurrence-free survival analysis on sub-cohorts. INR=Initial number at risk; FNR=Final (at 5yr) number at risk; RP=Radical prostatectomy.

|  | **Covariate** | **HR** | **95% CI** | | **p-value** |
| --- | --- | --- | --- | --- | --- |
| **Multivariable** | GG2 MRI (GG1 No MRI as reference) | 1.06 | 0.66 | 1.70 | 0.8 |
| **Univariate** | GG2 MRI (GG1 No MRI as reference) | 0.94 | 0.71 | 1.25 | 0.7 |

**Supplementary table 3.** Results of the sensitivity analysis of the recurrence-free survival analysis on with PSA and number of positive cores added as covariates.
